# Seroprevalence of Immunoglobulin G against measles and rubella over a 12-year period (2009 – 2021) in Kilifi, Kenya and the impact of the Measles-Rubella (MR) Vaccine campaign of 2016

**DOI:** 10.1101/2025.02.11.25322096

**Authors:** CN Mburu, J Ojal, R Selim, R Ombati, D Akech, B Karia, J Tuju, A Sigilai, G Smits, PGM van Gageldonk, FRM van der Klis, S Flasche, EW Kagucia, JAG Scott, IMO Adetifa

## Abstract

**Background:** Measles and rubella have been targeted for elimination by the World Health Organization. Age-specific population immunity to measles and rubella is important to assess progress towards elimination but data are scarce. We conducted seroprevalence surveys to identify disease-specific population immunity profiles in children and adults in Kilifi.

**Methods:** Sera from cross-sectional surveys in the Kilifi Health Demographic Surveillance System (2009–2021) were analysed using a fluorescent bead-based multiplex immunoassay. Bayesian multilevel regression with post stratification was used to obtain seroprevalence estimates adjusted for the underlying population and assay performance. Associations between seropositivity and age, sex, location and ethnic group were assessed using a mixed effects logistic regression.

**Results:** Measles-adjusted seroprevalence showed a significant increase from 88% in 2009 to 93% in 2021 (τ = 0.875, P = 0.01). Seropositivity was significantly higher in all age groups compared to those under 9 months. Seroprevalence among children ineligible for the first measles vaccine dose (MCV1) remained low (10–57%), whereas MCV1-eligible children (9–17 months) had higher seroprevalence (68–91%). Adult measles seroprevalence exceeded 96%. Rubella seroprevalence followed a similar pattern, with adults above 88%. Following the MR campaign, measles seroprevalence increased from 92% to 96% in eligible children, while rubella seroprevalence rose from 45% to 82%

**Conclusion:** Population immunity for measles significantly increased over the 12-year period suggesting improvement in immunisation program performance. To reduce reliance on frequent SIAs, efforts should focus on optimising both the timing and coverage of routine doses, particularly ensuring higher coverage of MCV2. The introduction of rubella vaccination has positively impacted immunity in children. Sustaining this immunity is essential to prevent potential gaps in older age groups, which could increase the risk of Congenital Rubella Syndrome (CRS) in infants

## Background

Seroepidemiology is an effective tool to monitor population immunity against vaccine-preventable diseases (VPDs). Estimates of population immunity, particularly age-specific immunity profiles, can be used to identify high-risk groups that may require targeted intervention. They can also be used to monitor declining antibody levels in vaccine recipients, assess the impact of different vaccination schedules, and to estimate effective vaccination coverage, in some settings [1–4]. Given the high burden of VPDs, particularly in LMICs, serosurveys are underexploited for assessing the impact of vaccination programmes and for informing vaccine policy [5].

The Kenya Expanded Programme on Immunisation (KEPI) was introduced in 1980 with an initial schedule that included one dose of a Measles-Containing Vaccine (MCV1) given at 9 months [6] The first supplementary immunisation activity (SIA) with MCV which was conducted in 2002, targeted children aged 9 months to 14 years. Since then, SIAs have been conducted periodically every 3-4 years among under 5-year-olds or under 15-year-olds to prevent measles outbreaks in the country. However, Kenya has continued to experience periodic outbreaks including a large outbreak in 2006 following a delay in implementing an SIA [7], a second due to unvaccinated refugees arriving from neighbouring countries between 2010-2011[8] and a third following the postponement of the 2020 SIA[9]. These outbreaks have been attributed to the accumulation of susceptible children caused by sub-optimal routine coverage of MCV1 (consistently below the elimination target of 95%) and to the low uptake of MCV2 (between 28% to 57% since its introduction) [10]. Clinical rubella surveillance in Kenya is poor, consequently, the burden of infection is not well documented and only a few outbreaks have been reported, including a 2014 outbreak in a rural area; rubella is endemic in Kenya and outbreaks are often underestimated [11].

Although Kenya has made significant progress towards measles and rubella elimination, it has yet to meet the milestones for measles control set by the World Health Assembly (WHA)[12]. A second dose of MCV(MCV2), given at 18 months of age, was introduced in 2013 to decrease Kenya’s reliance on SIAs for measles control[13]. In 2016, in response to a rise in rubella cases, a combined Measles-Rubella (MR) vaccine campaign was conducted in <15-year-olds with an achieved estimated coverage of 95% [14]. Thereafter the MR vaccine replaced monovalent measles vaccine in the routine immunisation schedule at 9 months and 18 months[15].

Data on population immunity against measles and rubella are limited in Kenya. The present study was undertaken to review annual trends (2009-2021) in age-specific population immunity, with the aim of identifying susceptible populations among both children (aged <15 years) and adults. In addition, we also assessed the impact of a single SIA by exploring changes in measles and rubella population immunity before and after the 2016 campaign.

## Methods

### Study Setting, study population and survey design

This study utilised a retrospective series of cross-sectional serum samples from the Kilifi Health & Demographic Surveillance System (KHDSS) population accessed from three prior serological studies in the KEMRI-Wellcome Trust Research Program (KWTRP) biobank. These observational studies included residents registered in KHDSS which monitors births, deaths, in-migration and out-migration in a population of over 300,000 of Kilifi County’s approximately 1.4 million residents [16].

Plasma or serum samples from 2009 to 2019 were obtained from three survey frameworks: (i) the Malaria Cross-Sectional Survey (2009-2013)[17], which was conducted annually as an independent, random, cross-sectional, sample from among a rolling cohort of healthy children resident in the KHDSS - since 1998, children have been recruited into this cohort continuously and followed until age 15; (ii) the Pneumococcal Conjugate Vaccine Impact Study i.e. PCVIS (2015-2019) [18],which conducted annual, cross-sectional surveys of KHDSS residents; (iii) the COVID-19 serosurveillance study conducted in 2021 in response to the pandemic sampling residents of the KHDSS at random [19]. Sera in adults (≥15 years old) were only available in this one study year, 2021.

The samples from 2009 to 2013 which were obtained from the Malaria Cross-sectional survey were a subset of the initial survey. This subset was randomly selected from each age group in the population register of the KHDSS in 2009, 2011 and 2013 in similar age-strata to the PCVIS samples, i.e., 10 age strata (0, 1, 2, 3, 4, 5, 6, 7, 8–9, and 10–14 years), with 50 children in each stratum[20]. We assayed sera collected from all participants in the PCVIS study (2015-2019) and the COVID-19 serosurveillance study (2021). Participants in the PCVIS study were selected using an independent age-stratified random selection method from the KHDSS population in 10 age strata (0, 1, 2, 3, 4, 5, 6, 7, 8–9, and 10–14 years) with 50 children in each stratum. For 2021, 100 individuals were randomly selected in each 5-year age band from 0-14 years, 50 individuals in each 5-year age band from 15-64 years and 50 individuals aged ≥ 65 years bringing the total to 850 participants; this was the complete set of the COVID-19 serosurveillance study samples in KHDSS. In the original studies, about 2mls of venous blood were collected from all consenting participants then separated, aliquoted, and stored at −70°C until processing.

### Bead coupling and fluorescent bead multiplex immunoassay

IgG antibodies against measles and rubella were determined using a fluorescent bead-based multiplex immunoassay on a Luminex^®^ MAGPIX^®^ analyzer. The measles virus lysate, Edmonston strain (Zeptometrix) and Rubella antigen (HPV-77), Native Protein (Genway) were coupled to fluorescent beads, and the immunoassay was performed as previously described [21]. Briefly, serum/plasma samples (2.5µl) were diluted in duplicate (1/200 and 1/4000) in assay buffer containing phosphate-buffered saline, 0.1% Tween 20, and 3% bovine serum albumin. The International Standard for anti-rubella immunoglobulin (RUBI-1-94), 1600 IU/ml from the National Institute of Biological Standards and Control (NIBSC) was used as a multivalent reference for this multiplex bead immunoassay. RUBI-1-94 has been previously calibrated against the international standard serum for measles (66/202; 5 IU/ml; WHO, NIBSC), it contains a measles-specific IgG concentration of 62.74 IU/ml. The standard was 3-fold serially diluted in 11 steps starting at 1:50 dilution in assay buffer. The anti-measles and anti-rubella IgG concentrations were determined by interpolating the median fluorescent intensity (MFI) on the respective reference standard curves and expressed as IU/ml. To monitor assay performance, high and low in-house serum controls, standards and blanks were included on each plate. Analyte-specific QC trend analysis was conducted for every plate assayed, and the linearity and recovery of the standards were checked to confirm they were within the acceptable range (80%-120%). Samples were also assayed in duplicate dilutions (1/200 and 1/4000), and the coefficients of variation (CVs) of the IgG concentrations were checked to ensure they were within the acceptable range (<20%)[21].

For measles, correlates of protection against infection have been shown to range between 0.12 IU/ml and 2 IU/ml depending on the assay[22] while for rubella IgG levels exceeding 10 IU/ml are considered protective against infection[23]. Based on our assay, participants were classified as seropositive and considered protected against infection when IgG concentrations were ≥0.12 IU/ml for measles and ≥10.0 IU/ml for rubella.

### Ethical considerations

Ethical approval was obtained from the Scientific and Ethics Review Unit (SERU) of the Kenya Medical Research Institute (Protocol SERU 3847). The serological samples were collected under SERU-approved protocols with a provision for storage of residual samples and use in future research (SERU 1433, 4085, 2887, 3149, 3426). Written informed consent was obtained from parents/legal guardians of all participants before sample collection.

### Statistical analysis

We first tabulated the seropositive results in each survey year based on the age strata at the time of the sample collection, sex, ethnic group and location in KHDSS and generated crude seroprevalence in these age groups.

We reclassified the population immunity in children in relation to the national immunisation schedule into 6 age groups per year for analysis as follows: 0-8m (ineligible for vaccination), 9-17m (eligible for MCV1), 18-29m (eligible for MCV2), 30-59m, 5-9yrs and 10-14 yrs. To analyse immunity in adults, we classified all surveys using the 5-year age bands defined in the 2021 survey.

During the study period (2009-2019), the age-specific seroprevalence showed no significant variation within individual-year age groups across the six defined age strata. Consequently, we did not calculate a summary weighted seroprevalence and instead grouped the data into these six age strata for analysis. To facilitate comparisons across years while removing the influence of age, we standardized seroprevalence using a reference population. We used the average population structure from the entire series as our standard population for each of the yearly samples to standardize both the age-specific seroprevalence values as well as the total population values. This standardisation was implemented using multilevel regression and post-stratification (MLRP) adjusted for sensitivity and specificity of the assay. The sensitivity of the measles and rubella assays was given as 96.5% and 99%, respectively, and the specificity was 95% and 95%, respectively[21]. The MLRP was implemented by fitting a Bayesian logistic regression that included age as a variable. Non-informative priors were used for all parameters and the models were fitted using the rjags software package[24].

Bayesian population-weighted and test-adjusted seroprevalence estimates were visualized using bar graphs and differences between groups in each year were tested using chi-square and Fisher’s exact tests where appropriate. We used a mixed-effects logistic regression to test for association of seroprevalence with year, sex, age, location and ethnic group.

Geometric mean concentrations (GMCs) and the 95% confidence levels were also calculated, and values were tabulated. Similar to the seroprevalence estimates, to facilitate comparisons across years while removing the influence of age, we used the average population structure from the entire series as our standard population for each of the yearly samples to standardize both the age-specific GMCs as well as the annual GMCs. One-way ANOVA was used to test differences in GMCs between groups in each year. Raw antibody titres were log-transformed and displayed via boxplots to show the variation within each age group across different years.

To assess the impact of the MR campaign carried out in May 2016, we identified the cohort of children aged between 9 months to 14 years at the time who were therefore eligible to receive the vaccine; these children were born between mid-May 2002 and mid-August 2015. We restricted and stratified the samples from these children into pre-campaign samples i.e., those collected in the 2015 survey, and post-campaign population i.e., those collected in the 2017 survey. To facilitate comparison in similar age groups, we excluded children aged 0-8m in the pre-campaign period as this age group did not have a corresponding group in the post-campaign period. To facilitate comparisons between the two periods while removing the influence of age, we used the average population structure from the two surveys (2015 and 2017) to standardize the age-specific GMCs as well as the annual GMCs. Seroprevalence estimates were also standardized in the same manner, and these were further adjusted for test sensitivity and specificity using MLRP. Differences pre- and post-campaign assessed using a chi-square test and paired sample t-test. All statistical analyses were conducted using the R-statistical software.

## Results

In total, there were 2,686 participants. The distribution of the samples by age, sex, location and ethnic group is shown in Table 1. Overall, there was a slightly higher number of samples from male participants (1381/2686, 51%). The majority of samples came from North and South locations in Kilifi and the Chonyi and Giriama ethnic groups. The number of samples assayed each year varied from 290 to 520.

**Table 1.** Crude measles (M) and rubella (R) seropositivity and total number of participants(N) per survey year stratified by age during sample collection, sex, location in KHDSS and ethnic group.

| Survey year | 2009<br>(5 June-19 Sep) |  |  | 2011<br>(25 Jun-4 Nov) |  |  | 2013<br>(29 Jun-10 Nov) |  |  | 2015<br>(1 Jul-31 Oct) |  |  | 2017<br>(28 Jun-8 Nov) |  |  | 2019<br>(29 Jun-7 Jul 2019) |  |  | 2021<br>(1 Jan-31 May) |  |  |  |
| --- | --- | --- | --- | --- | --- | --- | --- | --- | --- | --- | --- | --- | --- | --- | --- | --- | --- | --- | --- | --- | --- | --- |
|  | N | M(%) | R(%) | N | M(%) | R(%) | N | M(%) | R(%) | N | M(%) | R(%) | N | M(%) | R(%) | N | M(%) | R(%) |  | N | M(%) | R(%) |
| Age in years |  |  |  |  |  |  |  |  |  |  |  |  |  |  |  |  |  |  |  |  |  |  |
| <1 | 13 | 31 | 0 | 14 | 43 | 0 | 7 | 14 | 0 | 35 | 46 | 14 | 25 | 16 | 16 | 39 | 28 | 31 | 0-4 | 92 | 88 | 87 |
| 1 | 31 | 87 | 6 | 19 | 89 | 21 | 43 | 93 | 0 | 33 | 88 | 24 | 41 | 88 | 61 | 48 | 94 | 96 | 5_9 | 103 | 92 | 89 |
| 2 | 30 | 90 | 7 | 22 | 100 | 5 | 22 | 100 | 0 | 33 | 97 | 15 | 35 | 94 | 74 | 49 | 88 | 90 | 10_14 | 95 | 97 | 97 |
| 3 | 33 | 94 | 15 | 18 | 100 | 22 | 30 | 100 | 13 | 38 | 89 | 29 | 48 | 100 | 79 | 53 | 91 | 91 |  |  |  |  |
| 4 | 37 | 100 | 24 | 22 | 100 | 14 | 25 | 100 | 8 | 38 | 89 | 42 | 36 | 94 | 86 | 54 | 96 | 89 |  |  |  |  |
| 5 | 41 | 90 | 32 | 18 | 100 | 28 | 24 | 96 | 13 | 35 | 94 | 46 | 50 | 98 | 88 | 50 | 90 | 72 |  |  |  |  |
| 6 | 62 | 98 | 39 | 22 | 95 | 45 | 32 | 97 | 9 | 32 | 97 | 50 | 46 | 96 | 85 | 46 | 96 | 83 |  |  |  |  |
| 7 | 38 | 92 | 53 | 32 | 94 | 56 | 25 | 92 | 28 | 37 | 95 | 49 | 46 | 96 | 91 | 60 | 98 | 88 |  |  |  |  |
| 8-9 | 56 | 88 | 48 | 65 | 94 | 69 | 62 | 94 | 48 | 48 | 92 | 58 | 40 | 95 | 95 | 65 | 92 | 92 |  |  |  |  |
| 10-14 | 24 | 83 | 71 | 76 | 84 | 72 | 136 | 85 | 68 | 47 | 89 | 66 | 54 | 94 | 96 | 56 | 100 | 95 |  |  |  |  |
| Sex |  |  |  |  |  |  |  |  |  |  |  |  |  |  |  |  |  |  |  |  |  |  |
| Female | 179 | 95 | 32 | 151 | 96 | 50 | 197 | 99 | 33 | 172 | 93 | 40 | 212 | 96 | 83 | 262 | 94 | 86 |  | 132 | 99 | 95 |
| Male | 186 | 94 | 32 | 157 | 94 | 44 | 209 | 93 | 36 | 204 | 92 | 41 | 209 | 95 | 79 | 258 | 93 | 84 |  | 158 | 95 | 89 |
| Location |  |  |  |  |  |  |  |  |  |  |  |  |  |  |  |  |  |  |  |  |  |  |
| North | 83 | 95 | 15 | 2 | 99 | 0 | 90 | 97 | 32 | 150 | 91 | 31 | 158 | 96 | 84 | 157 | 93 | 82 |  | 2 | 100 | 99 |
| South | 206 | 98 | 37 | 251 | 95 | 46 | 230 | 96 | 40 | 108 | 94 | 44 | 130 | 91 | 81 | 187 | 93 | 90 |  | 2 | 51 | 50 |
| Township | 5 | 84 | 19 | 1 | 99 | 99 | 4 | 78 | 24 | 111 | 91 | 49 | 119 | 97 | 77 | 172 | 96 | 82 |  | 0 | 0 | 0 |
| Unspecified | 71 | 86 | 41 | 54 | 93 | 52 | 82 | 95 | 21 | 7 | 99 | 72 | 14 | 99 | 86 | 4 | 51 | 99 |  | 286 | 97 | 92 |
| Ethnic group |  |  |  |  |  |  |  |  |  |  |  |  |  |  |  |  |  |  |  |  |  |  |
| Chonyi | 129 | 93 | 35 | 144 | 93 | 42 | 138 | 90 | 34 | 127 | 98 | 43 | 135 | 92 | 80 | 184 | 95 | 89 |  | 2 | 51 | 50 |
| Giriama | 105 | 98 | 19 | 34 | 96 | 44 | 112 | 99 | 36 | 173 | 87 | 38 | 195 | 95 | 81 | 222 | 94 | 84 |  | 2 | 100 | 99 |
| Jibana | 48 | 99 | 44 | 57 | 99 | 56 | 58 | 96 | 45 | 5 | 99 | 40 | 7 | 90 | 86 | 9 | 81 | 78 |  | 0 | 0 | 0 |
| Kauma | 7 | 99 | 0 | 12 | 96 | 59 | 12 | 99 | 50 | 48 | 92 | 35 | 44 | 99 | 82 | 72 | 95 | 78 |  | 0 | 0 | 0 |
| Others | 76 | 85 | 41 | 61 | 93 | 49 | 86 | 95 | 23 | 23 | 96 | 61 | 40 | 97 | 83 | 33 | 82 | 82 |  | 286 | 97 | 92 |
| Total | 365 | 90 | 40 | 308 | 91 | 46 | 406 | 91 | 33 | 376 | 88 | 47 | 421 | 90 | 82 | 520 | 89 | 86 |  | 290 | 92 | 90 |

The overall Bayesian age-standardised proportion of children with protective measles antibodies in Kilifi varied between 88% CI: (81-92%) in 2009 to 93% CI: (90-97%) in 2021 (Table 2a, 2b and Fig. 1a). A Mann-Kendall trend test indicated a significant increasing trend in measles seroprevalence over the 12-year period (*tau*= 0.875, P = 0.01). In all the surveys, there was significant heterogeneity in seroprevalence by age (P<0.05). This pattern was consistent over time; MCV1 ineligible children always had the lowest seroprevalence ranging between 10-57%; seroprevalence was consistently higher in the MCV1 eligible children (68%-91%); and seroprevalence continued to increase in MCV2 eligible children(18-29m). Children in the 5-9- and 10-14-year age groups had a seroprevalence estimate of >95% in all study years.

**Figure 1.**
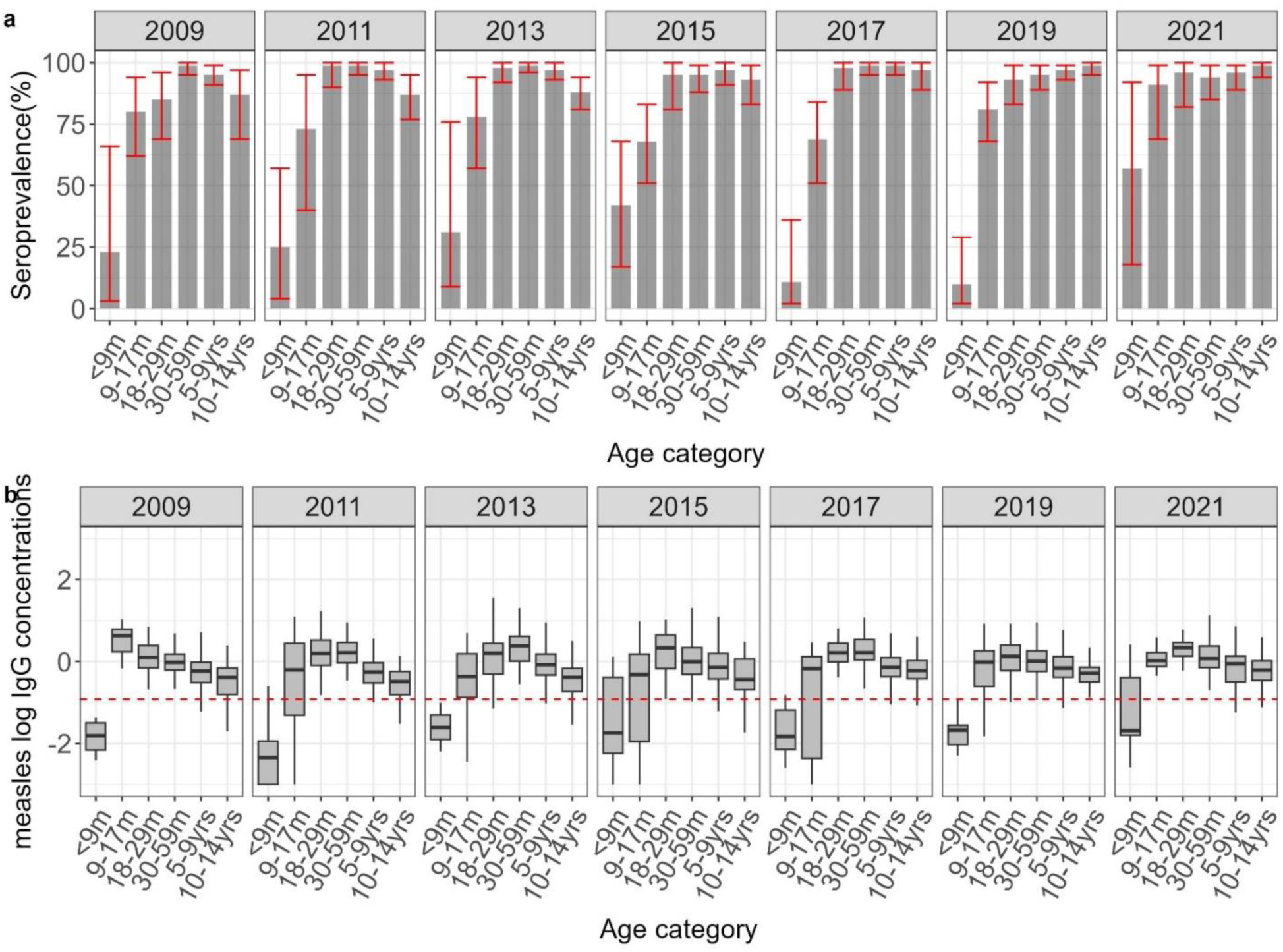
1a shows standardized age-specific measles seroprevalence estimates adjusted for test performance using Bayesian multilevel regression and poststratification. The red error bars indicate 95% credible intervals. 1b shows the distribution of antibodies with the median indicated by the line in the box and a whisker plot. The red line in the lower figure is the log of the protective threshold for measles of ≥0.12 IU/ml.

**Table 2a.** Crude and population-weighted and test-performance adjusted measles and rubella IgG seroprevalence estimates by survey year (2009,2011,2013,2015). Seroprevalence estimates were calculated by using multilevel regression and poststratification (MLRP) to account for differences in population across the years and subsequently adjusted for assay sensitivity and specificity.

| Survey year | 2009 |  |  |  |  | 2011 |  |  |  |  | 2013 |  |  |  |  | 2015 |  |  |  |  |
| --- | --- | --- | --- | --- | --- | --- | --- | --- | --- | --- | --- | --- | --- | --- | --- | --- | --- | --- | --- | --- |
|  |  | Crude seroprevalence |  | Bayesian population-weighted and test-adjusted |  |  | Crude seroprevalence |  | Bayesian population-weighted and test-adjusted |  |  | Crude seroprevalence |  | Bayesian population-weighted and test-adjusted |  |  | Crude seroprevalence |  | Bayesian population-weighted and test-adjusted |  |
| measles | N | % [95% CI] |  | % [95% CI] |  | N | % [95% CI] |  | % [95% CI] |  | N | % [95% CI] |  | % [95% CI] |  | N | % [95% CI] |  | % [95% CI] |  |
| Age |  |  |  |  |  |  |  |  |  |  |  |  |  |  |  |  |  |  |  |  |
| <9m | 4 | 0 | [0-49] | 23 | [3-66] | 8 | 13 | [2-47] | 25 | [4-57] | 3 | 0 | [0-56] | 31 | [9-76] | 14 | 36 | [16-61] | 42 | [17-68] |
| 9-17m | 26 | 77 | [58-89] | 80 | [62-94] | 9 | 67 | [35-88] | 73 | [40-95] | 19 | 74 | [51-88] | 78 | [57-94] | 35 | 66 | [49-79] | 68 | [51-83] |
| 18-29m | 34 | 82 | [66-92] | 85 | [69-96] | 24 | 100 | [86-100] | 99 | [90-100] | 38 | 97 | [87-100] | 98 | [92-100] | 31 | 90 | [75-97] | 95 | [81-100] |
| 30-59m | 80 | 98 | [91-99] | 99 | [95-100] | 54 | 100 | [93-100] | 99 | [95-100] | 67 | 100 | [95-100] | 99 | [96-100] | 97 | 92 | [85-96] | 95 | [88-99] |
| 5-9yrs | 197 | 92 | [88-95] | 95 | [91-99] | 137 | 95 | [90-98] | 97 | [93-100] | 143 | 94 | [89-97] | 97 | [93-100] | 152 | 94 | [89-97] | 97 | [91-100] |
| 10-14yrs | 24 | 83 | [64-93] | 87 | [69-97] | 76 | 84 | [74-91] | 87 | [77-95] | 136 | 85 | [78-90] | 88 | [81-94] | 47 | 89 | [77-95] | 93 | [83-99] |
| Total | 365 | 90 | [86-93] | 88 | [81-92] | 308 | 91 | [87-93] | 89 | [85-93] | 406 | 91 | [88-93] | 90 | [86-93] | 376 | 88 | [84-91] | 90 | [86-94] |
| P value |  | <0.001 |  | <0.001 |  |  | <0.001 |  | <0.001 |  |  | <0.001 |  | <0.001 |  |  | <0.001 |  | <0.001 |  |
| rubella |  |  |  |  |  |  |  |  |  |  |  |  |  |  |  |  |  |  |  |  |
| <9m | 4 | 0 | [0-49] | 11 | [1-50] | 8 | 0 | [0-32] | 6 | [0-31] | 3 | 0 | [0-56] | 13 | [1-59] | 14 | 21 | [8-48] | 22 | [5-47] |
| 9-17m | 26 | 0 | [0-13] | 3 | [0-14] | 9 | 22 | [6-55] | 24 | [4-57] | 19 | 0 | [0-17] | 3 | [0-15] | 35 | 17 | [8-33] | 17 | [6-33] |
| 18-29m | 34 | 6 | [2-19] | 6 | [1-18] | 24 | 13 | [4-31] | 12 | [3-29] | 38 | 0 | [0-9] | 2 | [0-9] | 31 | 16 | [7-33] | 16 | [6-33] |
| 30-59m | 80 | 20 | [13-30] | 19 | [11-30] | 54 | 13 | [6-24] | 13 | [5-25] | 67 | 9 | [4-18] | 8 | [2-17] | 97 | 32 | [24-42] | 32 | [23-42] |
| 5-9yrs | 197 | 43 | [36-50] | 44 | [36-51] | 137 | 57 | [49-65] | 59 | [50-67] | 143 | 30 | [23-38] | 30 | [23-39] | 152 | 51 | [43-59] | 53 | [44-62] |
| 10-14yrs | 24 | 71 | [51-85] | 74 | [53-89] | 76 | 72 | [61-81] | 75 | [63-85] | 136 | 68 | [59-75] | 70 | [61-78] | 47 | 66 | [52-78] | 68 | [53-81] |
| Total | 365 | 33 | [28-38] | 42 | [34-49] | 308 | 47 | [42-53] | 48 | [42-53] | 406 | 35 | [30-39] | 34 | [30-39] | 376 | 41 | [36-46] | 48 | [41-54] |
| P value |  | <0.001 |  | <0.001 |  |  | <0.001 |  | <0.001 |  |  | <0.001 |  | <0.001 |  |  | <0.001 |  | <0.001 |  |

**Table 2b.** Crude and population-weighted and test-performance adjusted measles and rubella IgG seroprevalence estimates by survey year (2017,2019,2021). Seroprevalence estimates were calculated by using multilevel regression and poststratification (MLRP) to account for differences in population across the years and subsequently adjusted for assay sensitivity and specificity.

| Survey year | 2017 |  |  |  |  | 2019 |  |  |  |  | 2021 |  |  |  |  |
| --- | --- | --- | --- | --- | --- | --- | --- | --- | --- | --- | --- | --- | --- | --- | --- |
|  |  | Crude seroprevalence |  | Bayesian population-weighted and test-adjusted |  |  | Crude seroprevalence |  | Bayesian population-weighted and test-adjusted |  |  | Crude seroprevalence |  | Bayesian population-weighted and test-adjusted |  |
| measles | N | % [95% CI] |  | % [95% CI] |  | N | % [95% CI] |  | % [95% CI] |  | N | % [95% CI] |  | % [95% CI] |  |
| Age |  |  |  |  |  |  |  |  |  |  |  |  |  |  |  |
| <9m | 15 | 7 | [1-30] | 11 | [2-36] | 20 | 5 | [1-24] | 10 | [2-29] | 5 | 40 | [12-77] | 57 | [18-92] |
| 9-17m | 36 | 67 | [50-80] | 69 | [51-84] | 46 | 78 | [64-88] | 81 | [68-92] | 15 | 87 | [62-96] | 91 | [69-99] |
| 18-29m | 34 | 97 | [85-99] | 98 | [89-100] | 56 | 89 | [79-95] | 93 | [83-99] | 20 | 95 | [76-99] | 96 | [82-100] |
| 30-59m | 100 | 97 | [92-99] | 99 | [95-100] | 121 | 93 | [86-96] | 95 | [89-99] | 52 | 90 | [79-96] | 94 | [85-99] |
| 5-9yrs | 182 | 96 | [92-98] | 99 | [95-100] | 221 | 94 | [90-97] | 97 | [93-99] | 103 | 92 | [85-96] | 96 | [89-99] |
| 10-14yrs | 54 | 94 | [85-98] | 97 | [89-100] | 56 | 100 | [94-100] | 99 | [95-100] | 95 | 97 | [91-99] | 99 | [94-100] |
| Total | 421 | 90 | [87-93] | 92 | [88-94] | 520 | 89 | [86-91] | 91 | [89-93] | 290 | 92 | [89-95] | 93 | [90-97] |
| P value |  | <0.001 |  | <0.001 |  |  | <0.001 |  | <0.001 |  |  | 0.01 |  | 0.01 |  |
| rubella |  |  |  |  |  |  |  |  |  |  |  |  |  |  |  |
| <9m | 15 | 7 | [1-30] | 8 | [1-28] | 20 | 10 | [3-30] | 11 | [2-28] | 5 | 60 | [23-88] | 62 | [20-93] |
| 9-17m | 36 | 58 | [42-73] | 60 | [42-76] | 46 | 78 | [64-88] | 81 | [66-92] | 15 | 87 | [62-96] | 88 | [66-99] |
| 18-29m | 34 | 65 | [48-79] | 67 | [49-81] | 56 | 91 | [81-96] | 94 | [85-99] | 20 | 95 | [76-99] | 95 | [80-100] |
| 30-59m | 100 | 80 | [71-87] | 83 | [74-91] | 121 | 90 | [83-94] | 93 | [86-99] | 52 | 87 | [75-93] | 89 | [78-98] |
| 5-9yrs | 182 | 90 | [84-93] | 93 | [87-98] | 221 | 85 | [79-89] | 88 | [81-93] | 103 | 89 | [82-94] | 93 | [85-99] |
| 10-14yrs | 54 | 96 | [87-99] | 97 | [90-100] | 56 | 95 | [85-98] | 96 | [89-100] | 95 | 97 | [91-99] | 98 | [94-100] |
| Total | 421 | 81 | [76-84] | 84 | [80-87] | 520 | 84 | [81-87] | 87 | [83-90] | 290 | 91 | [87-94] | 91 | [87-96] |
| P value |  | <0.001 |  | <0.001 |  |  | <0.001 |  | <0.001 |  |  | 0.03 |  | 0.03 |  |

GMC levels varied significantly across the ages (P <0.05) in all the surveys (Table s3) although there was no consistent trend observed across the years. From the distribution of the log of concentration plots, the median showed a declining trend with increasing age (Fig. 1b). Evidence of waning maternal antibodies was seen in MCV1 ineligible children, reflected by low median of log antibody concentrations. Furthermore, waning vaccine-induced immunity was apparent from the decline in median of log antibody levels across age groups, with some older age groups in specific survey years nearing the threshold (Fig.1b). Increased immunity following the three SIAs during our study period was evident in both the log antibody distribution figures and GMCs table, particularly in surveys conducted after the SIAs in 2009, 2012, and 2016.

In the mixed-effects logistic regression, children in the age groups 9-17 months, 18-29 months, 30-59 months, 5-9 years, and 10-14 years had significantly higher measles seropositivity compared to children aged <9 months (95% CI did not include 1). Additionally, children from the Jibana ethnic group showed significantly higher measles seropositivity compared to the Chonyi ethnic group (OR = 1.91, 95% CI = 1.62-15.98) and females exhibited significantly higher measles seropositivity compared to males(OR = 1.42, 95% CI = 1.06-1.89) (Table 3).

**Table 3.**
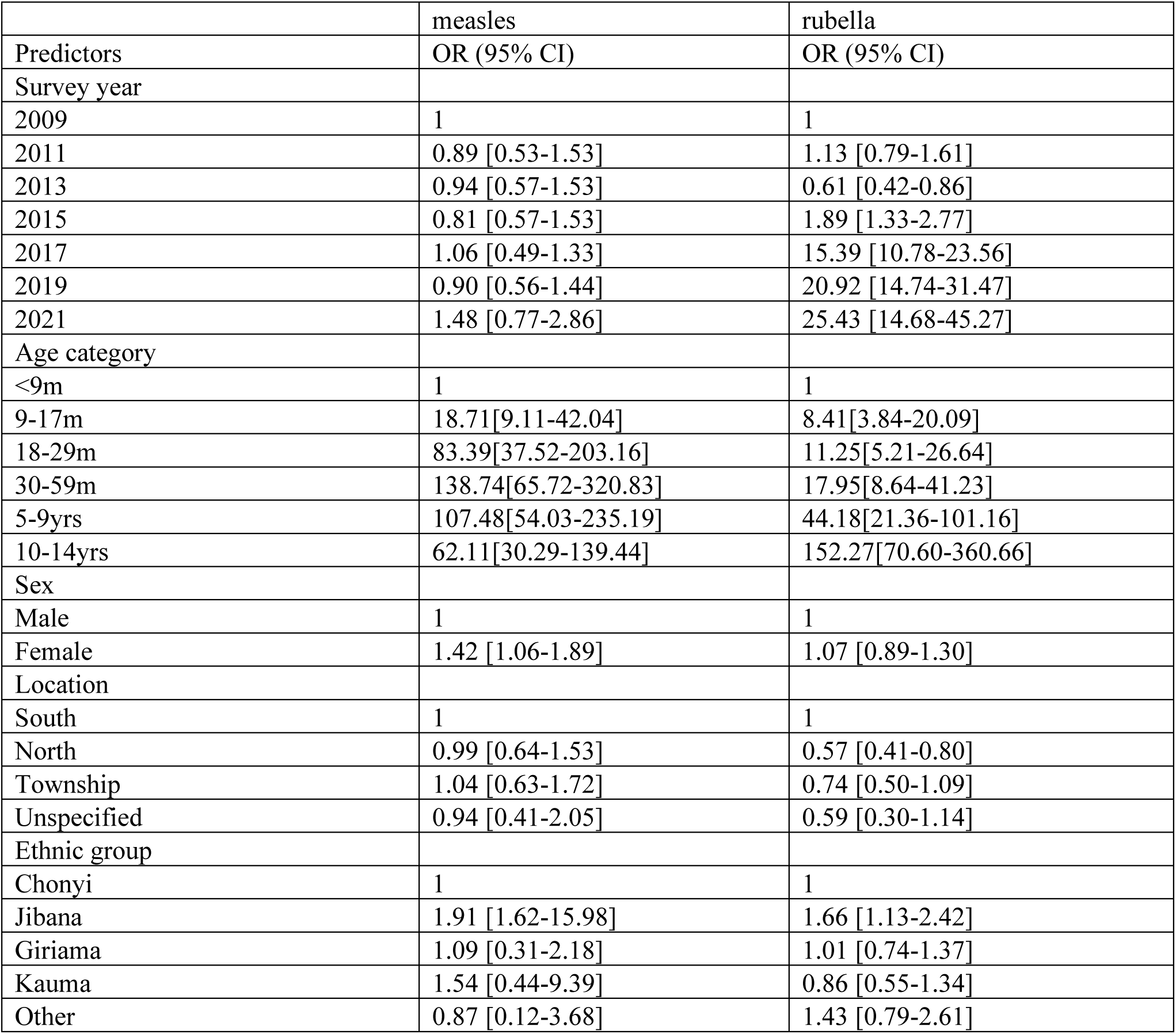
shows the multivariable factors associated with measles and rubella seroprevalence. The analysis was based on the raw estimates presented in table 1. The explanatory factors included survey year (with 2009 as the reference), age category (with <9m as the reference category), sex (with male as the reference category), location (with south as the reference) and ethnic group (with Chonyi as the reference).

Measles seroprevalence in adults was high ranging between 96-100% and comparable across the different ages (P=0.41, Table s5). GMCs in adults were high and increased across the ages (Table s5). Distribution of log of concentration also showed an increasing trend (Fig.2)

**Figure 2.**
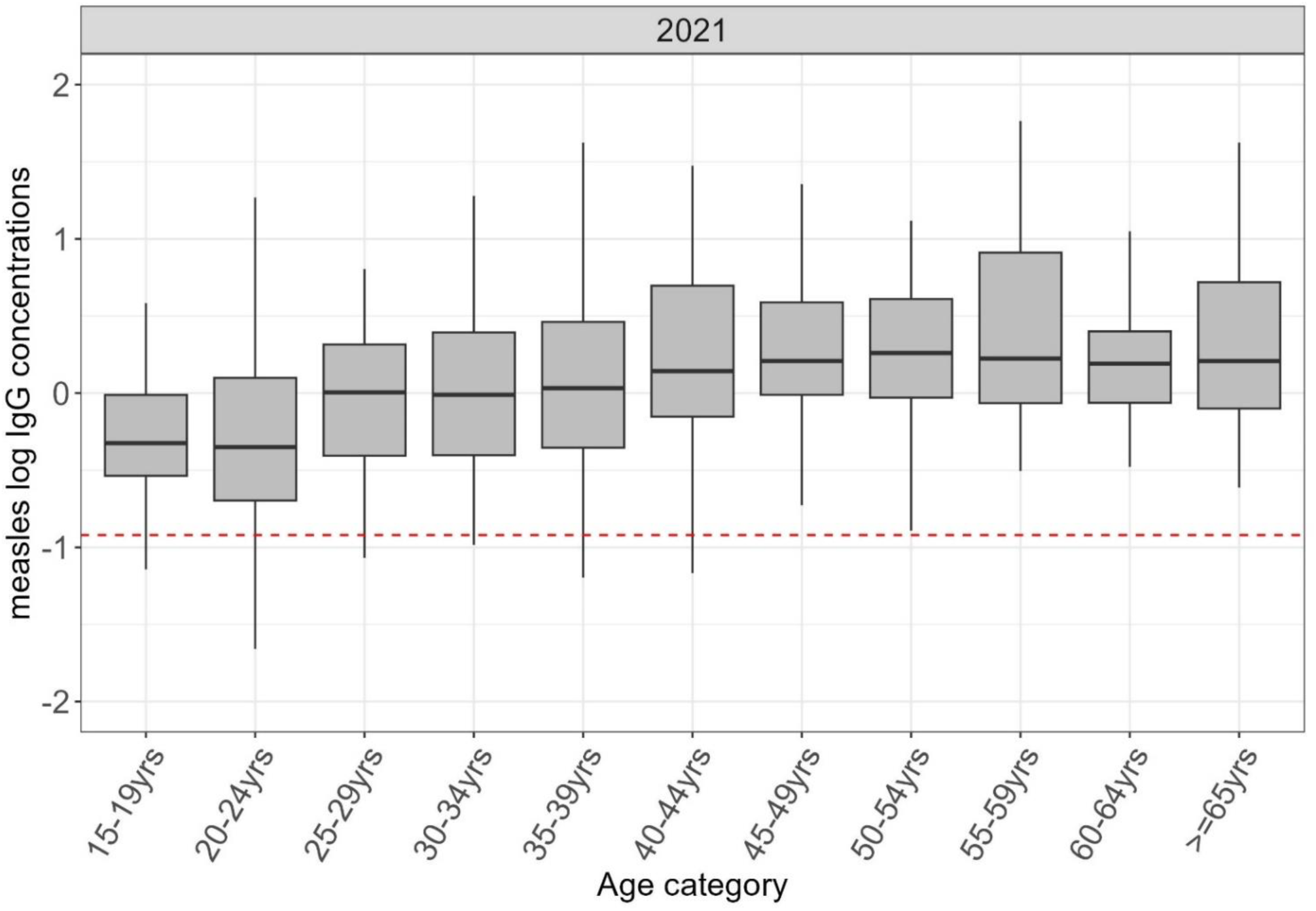
shows the distribution of log of measles antibodies in adults in KHDSS in 2021 with the median indicated by the line in the box and a whisker plot. The red line is the log of the protective threshold for measles of ≥0.12 IU/ml.

Prior to the introduction of the rubella vaccine in 2016, total seroprevalence among children under 15 years old ranged from 34% to 48% (Table 2a). After the vaccine was introduced, seroprevalence increased to between 82% and 90% (Table 2b and Fig. 3). Seroprevalence showed significant variation by age across all study years, as indicated by a chi-square test of proportions (P < 0.05).

**Figure 3.**
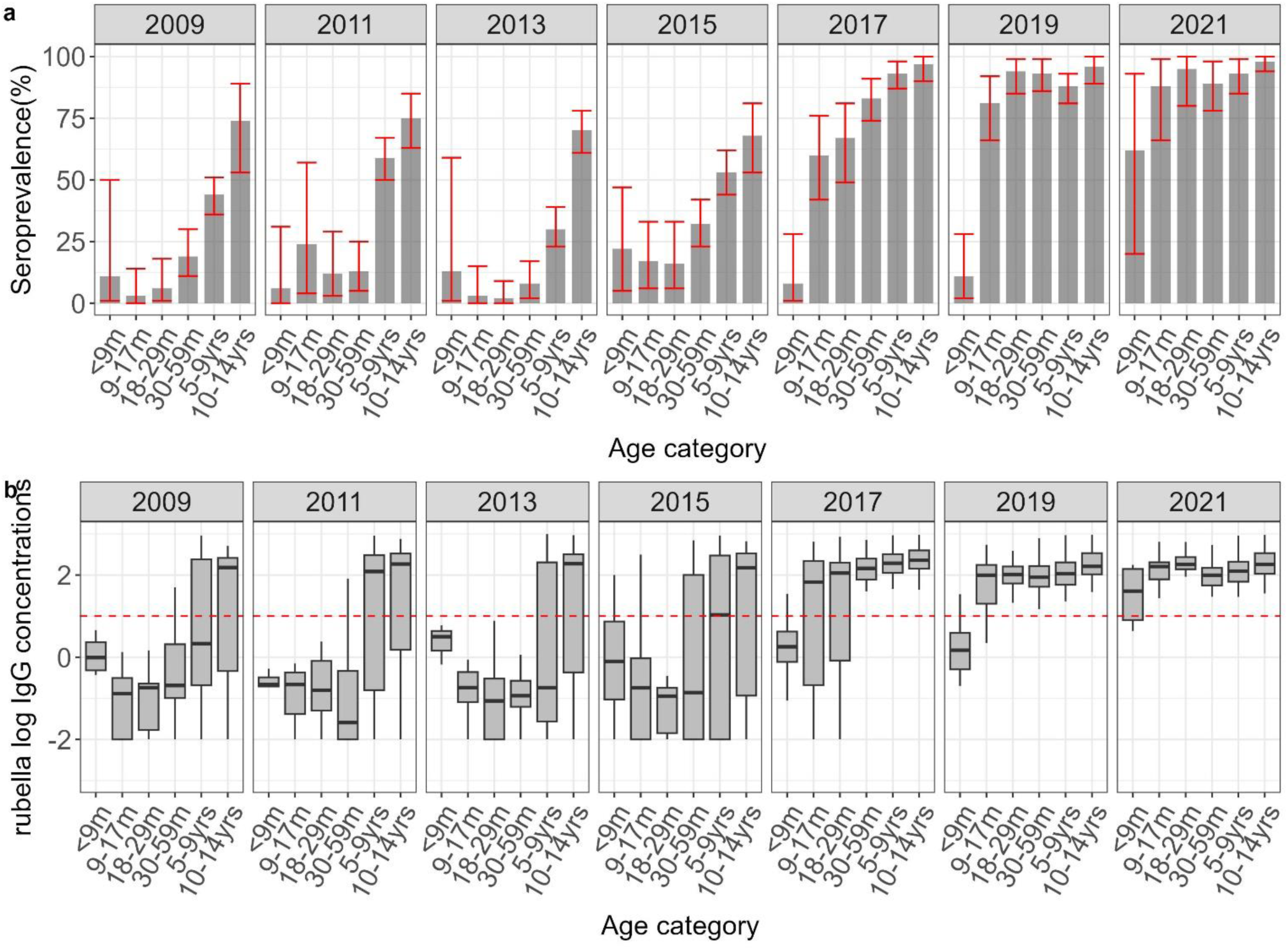
3a shows standardized age-specific rubella seroprevalence estimates adjusted for test performance using Bayesian multilevel regression and poststratification. The red error bars indicate 95% credible intervals. 3b shows the distribution of antibodies with the median indicated by the line in the box and a whisker plot. The red line in the lower figure is the log of the protective threshold for rubella of ≥10 IU/ml.

The pattern of rubella seroprevalence with age was similar in all survey years before vaccine introduction. Seroprevalence in the MR1 ineligible group ranged between 6% and 22% and seroprevalence in the MR1 eligible group ranged between 3% and 24%; from the second year of life, seroprevalence increased consistently with age.Following vaccine introduction, rubella seroprevalence increased more rapidly with age. MR1 ineligible children had the lowest seroprevalence, ranging between 8% and 62%. Seroprevalence increased in the MR1 eligible children and varied between 60% and 88% and, similarly, in MR2 eligible children, ranging between 67% and 95%. Seroprevalence increased among older age groups; in all the survey years seroprevalence was greater than 83% among older age groups (Table 2). GMC levels also increased significantly with age in all survey years (P <0.05, Table s4).

In the mixed-effects logistic regression, children aged 9-17 months, 18-29 months, 30-59 months, 5-9 years, and 10-14 years had significantly higher rubella seropositivity compared to children aged <9 months (95% CI excluded 1). Rubella seroprevalence did not show significant variation by sex, location, or ethnic group. However, seroprevalence in the years 2015, 2017, 2019, and 2021 was significantly higher compared to 2009 (Table 3).

Rubella seroprevalence in adults ranged between 88-99% and did not vary with age (p=0.91, Table s6). The distribution of log of antibody concentrations was comparable across the ages (Fig. 4)

**Figure 4.**
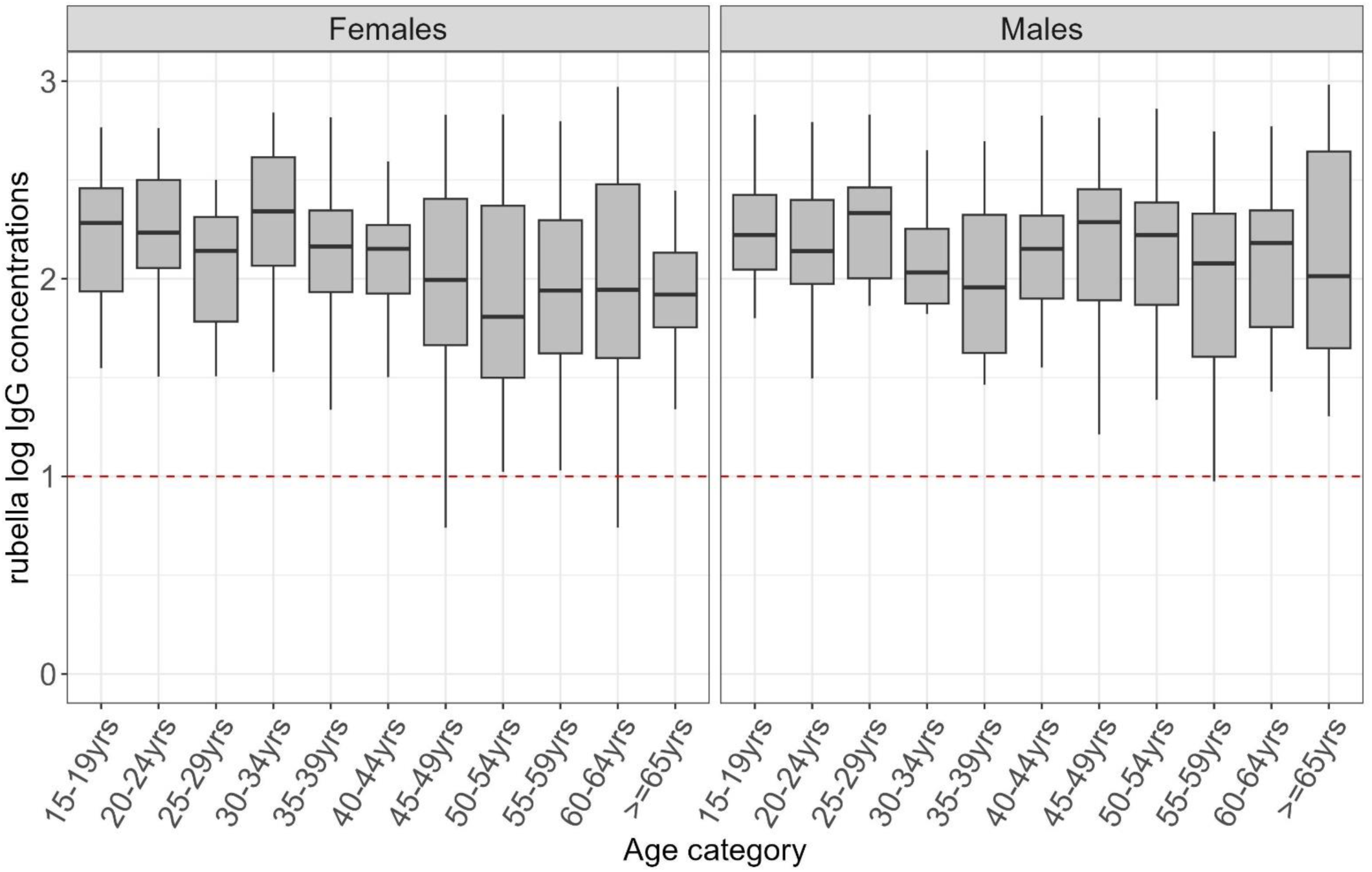
shows the distribution of antibodies to rubella in adults in KHDSS in 2021with the median indicated by the line in the box and a whisker plot. The red line in the lower figure is the log of the protective threshold for rubella of ≥10 IU/ml.

The pre- and post-campaign samples included sera from 351 and 370 children, respectively (Table s7). Measles seroprevalence was 92% (95% CI: 85-94%) in the pre-campaign period and showed a significant increase to 96% (95% CI: 89-98%) in the post-campaign period, as evidenced by a chi-square test (P = 0.01). Age-specific seroprevalence increased as follows: from 71% to 96% in children aged 9-17 months, from 95% to 98% in children aged 18-29 months, from 96% to 98% in children aged 30-59 months, from 97% to 98% in 5–9-year-olds, and from 94% to 97% in 10– 14-year-olds. This increase in seroprevalence was not significant in any age stratum (P>0.05). Overall, GMC levels increased significantly from 0.58 IU/ml (95% CI; 0.37-0.91 IU/ml) to 0.89 IU/ml (95% CI; 0.60-1.17 IU/ml) after the campaign (p<0.05). (Table s7 and Fig 5).

**Figure 5.**
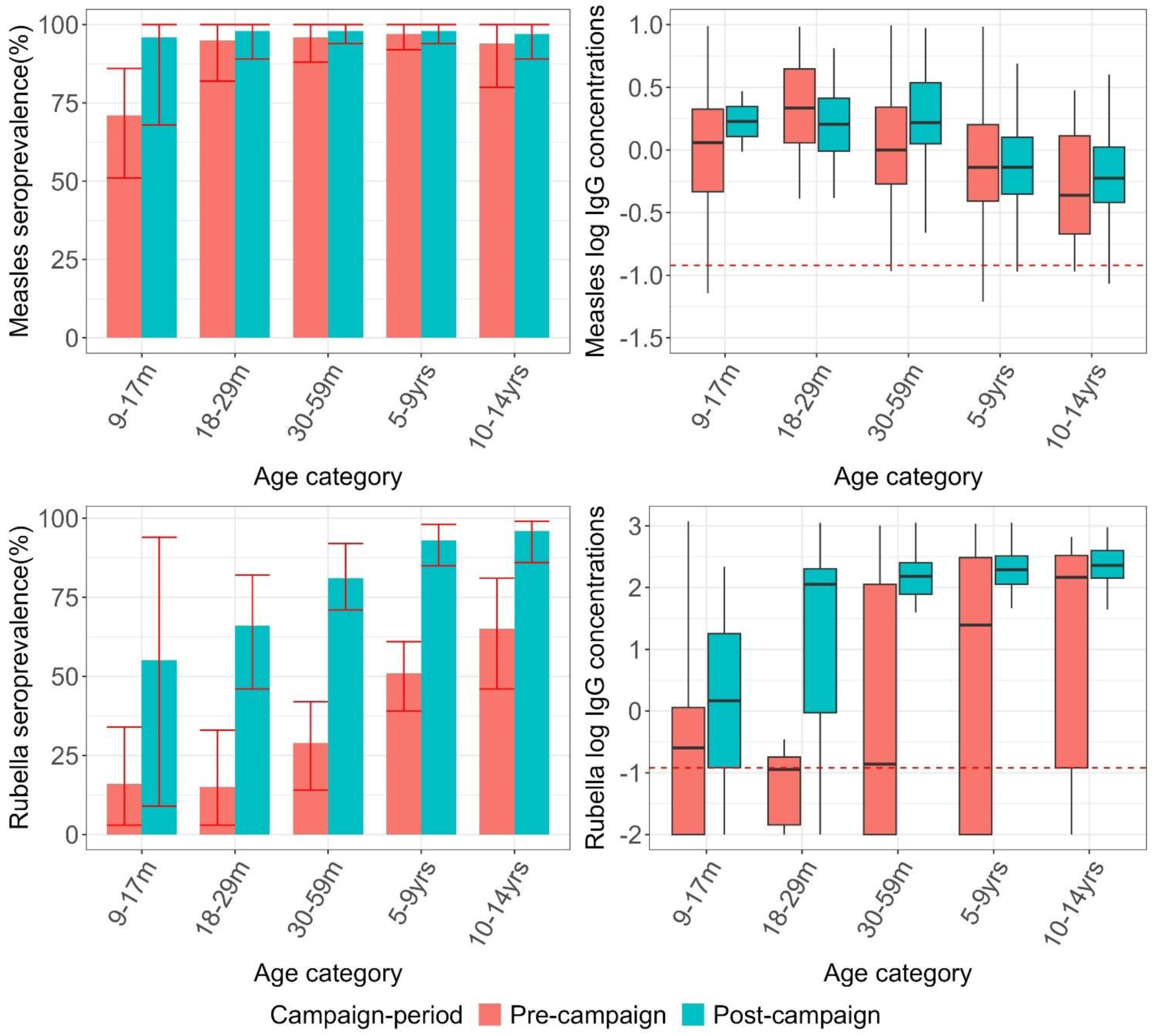
Age-specific measles and rubella seroprevalence and the distribution of antibodies with the median indicated by the line in the box and a whisker plot before and after the MR vaccine campaign of 2016.The red bars represent the seroprevalence in pre-campaign period while the green bars represent seroprevalence in the post-campaign in the age-eligible children.

The overall rubella seroprevalence among children aged 9 months to 14 years was estimated to be 45% (CI: 35-53%) before the campaign, and this increased significantly to 82% (CI: 77-88%) after the campaign period (chi-square test, P<0.001). Age-specific seroprevalence increased as follows: from 16% to 55% in children aged 9-17 months, from 15% to 66% in children aged 18-29 months, from 29% to 81% in children aged 30-59 months, from 51% to 93% in 5–9-year-olds, and from 65% to 96% in 10–14-year-olds. (Fig. 5, Table s7). Total population GMCs increased significantly from 4.8 IU/ml (95%CI:1.17-16.32) to 93.47 IU/ml (95% CI 58.50-151.98).

## Discussion

From 2009 to 2021, the overall proportion of children aged 0-14 years with protective measles antibodies in Kilifi exhibited a significant increasing trend over the 12-year period, ranging from 88% in 2009 to 93% in 2021. Although annual seroprevalence in most years remained below the herd immunity threshold for measles of 93%[25] indicating insufficient protection against outbreaks, the increasing seroprevalence in the last few years suggests ongoing improvement in KEPI performance over time. There is still some reliance on SIAs for boosting immunity, as evidenced by increased IgG antibody concentrations in older age groups following repeated SIAs, particularly in surveys conducted after these campaigns. As a result, despite antibody waning over time, a substantial proportion remains above the protection threshold, particularly in older age groups. For rubella, the results demonstrated the success of the vaccination programme; seroprevalence increased from 45% to 82% post-vaccine introduction in children and adults had a high seroprevalence of 92% in 2021. Sustaining robust coverage will be crucial to prevent immunity gaps in women of reproductive age and CRS in infants.

Children too young to have received MCV1 had a seroprevalence lower than 50% with fluctuations across the years. These findings indicate a prolonged period of susceptibility in young infants probably as a consequence of the rapid decay of maternally acquired antibodies. However, the small sample sizes in this age group, ranging from 4 to 20 children in each year, limit the ability to draw meaningful conclusions from this dataset. Additionally, upon closer examination of the monthly age distribution, we found that 69% of these children were aged 6 months or older. The rapid decay of maternal antibodies has been reported elsewhere[26, 27] particularly in areas where maternal immunity is from immunisation rather than natural infection[28]. The recommended age for MCV1 receipt is 9 months in high transmission settings[29]. Delaying infant vaccination aims to reduce interference from maternal antibodies, allowing optimal vaccine efficacy. However, the rate of maternal antibody waning varies, posing challenges in predicting the ideal vaccination timing. Early vaccination has been suggested in high-risk settings, although this is an ongoing discussion as moderate evidence indicates potential negative impacts on seroconversion to subsequent measles vaccine doses[30].

Seroprevalence in the MCV1 eligible group ranged from 68% to 91% across the years, reflecting variations likely influenced by differences in MCV1 uptake. Comparisons with reported administrative coverage of MCV1 during the same period, which ranged from 75% to 85%[31] also showed slight discrepancies. In some years, the reported coverage exceeded seroprevalence levels, while in others, it was lower. These discrepancies could be caused by several factors. Firstly, receiving a vaccine does not always result in immunisation against the targeted disease. Depending on the vaccine and the number of doses a minority of individuals may not develop an immune response post-vaccination. For example, following measles vaccination, the proportions of children who develop protective antibody levels are approximately 85% at 9 months of age and 95% at 12 months of age[32]. Secondly, administrative coverage estimates are prone to inaccuracies including errors in recording vaccine doses and incorrect assumptions about the target population’s size[33]. Thirdly, these discrepancies could also suggest poor timeliness of MCV1 vaccination which was previously reported across 6 different birth-cohorts in the KHDSS area[34]. The findings are consistent with those of several other studies that show the challenges of relying solely on vaccination coverage estimates as one might overlook these pockets of susceptibility which lead to outbreaks[1, 35].

Seroprevalence in the MCV2 eligible group remained consistently high both before and after introduction of the 2^nd^ dose. It is challenging to isolate the specific impact of MCV2 alone because this age group has benefited from repeated SIAs over the years. The lack of a noticeable change in seroprevalence could also be due to the low uptake of the second dose (range of 28-57%) since its introduction[31]. This low uptake, which has been reported in other areas in Africa, is influenced by factors such as knowledge, perceptions, and attitudes of the 2^nd^ dose at individual and community levels[36]. In addition, the dose timing, determined programmatically, does not align with the schedule of other RI visits which could also contribute to the low uptake[29]. Our findings reinforce the need for efforts to enhance awareness and improve the uptake of MCV2.

Seroprevalence in the older age groups was high as a result of repeated SIAs conducted over the years highlighting the program’s reliance on SIAs to maintain immunity levels. However, given WHO’s concerns about SIA sustainability in LMICs[29, 37], efforts should be made to improve the coverage of both MCV1 and 2 in the young age groups so as to sustain high seroprevalence levels in the older ages without the need for frequent SIA campaigns. Conducting a comprehensive analysis of trade-offs in the different intervention programs will be essential to identify optimal approaches for reducing reliance on these frequent and expensive SIAs.

In our analysis of the MR campaign of 2016, overall measles seroprevalence increased from 92-96%. The high seroprevalence prior to the campaign is likely due to the fact that children in the campaign target group would have benefited from the previous SIAs in 2009 and 2012[13]. A previous analysis looking at the impact of the 2002 measles vaccine campaign estimated an overall reduction in the susceptible population of about 65%[39]. The wide disparity between our estimates and the programmatic evaluation estimates is mostly driven by the fact that the pre-campaign population in the evaluation had relatively low immunity compared to our population. The pre- and post-campaign surveys in the evaluation were conducted one month apart whereas our pre- and post-campaign data were collected about 2 years apart.

Our findings illustrate the large impact on population immunity to rubella of the MR campaign and the introduction of routine rubella vaccination. Overall seroprevalence in children increased significantly from 45% to 82% post-campaign. Both the age-specific seroprevalence estimates and GMCs also increased significantly after the campaign. These findings are comparable to a similar study conducted in Zambia where serosurveys conducted before and after the introduction of the rubella vaccine demonstrated a significant increase in rubella seroprevalence from 51.3% (95% CI; 45.6-57.0%) to 98.3% (95% CI; 95.5-99.4%) in the campaign target group[40].

Age was positively associated with rubella seroprevalence even before the introduction of the vaccine as a result of the cumulative risk of natural infection with age[41]. The post-campaign seroprevalence in children indicates sufficient immunity levels to prevent outbreaks. However, sustaining high vaccination coverage remains essential for long-term protection, particularly in adults. Vaccinating young children may shift immunity dynamics, potentially reducing the circulation of wild-type rubella virus. Consequently, this could increase the susceptibility of older individuals to rubella if they are re-exposed. This is especially concerning for rubella, as infections in women of reproductive age can result in CRS in infants, leading to significant disabilities or even fatal outcomes[42]. Strengthening coverage is essential to control this risk and ensure comprehensive protection across all age groups.

This extensive seroprevalence study retrospectively utilised a series of cross-sectional surveys in KHDSS employing a random sampling strategy to ensure representativeness of children in the area. Testing was conducted using plasma or serum separated from venous blood, which, although more invasive than oral fluid samples[43] and dried blood spots[44], have demonstrated higher sensitivity[45]. Serum samples were simultaneously tested using a highly sensitive and specific fluorescent bead-based multiplex immunoassay[21] which has been shown to be propitious for sero-diagnostics for surveillance and epidemiology[46]. Additionally, the dataset was derived from a series of cross-sectional surveys conducted over different years, providing a temporal perspective on the evolving seroprevalence of measles and rubella across different age groups within the population.

The main limitation of this study lies in the generalisability of these findings to diverse settings. The study was carried out in a rural area within the African region, where most VPDs including measles and rubella are endemic. Although these results are representative of rural areas in measles and rubella endemic settings with similar vaccination coverage, the estimates may exhibit significant variation across rural and urban settings primarily due to differences in vaccination coverage, levels of natural exposure, and distinct mixing patterns across various age groups. Samples were also collected from individuals who agreed to participate and those who declined consent may have a different immunity profile.

The survey data were also from three distinct primary cross-sectional surveys conducted over a 12-year period. It is important to acknowledge that the extended freezing duration could potentially have led to antibody degradation in older samples which would generate a false impression of improved population immunity over time. In testing the serum samples, control standards were consistently applied on every plate to manage errors between plates and the samples were assayed in duplicate, and the coefficient of variation was monitored to ensure it remained below the acceptable threshold of <20%. The diagnostic accuracy of multiplex immunoassays, including both specificity and sensitivity, significantly impacts the resulting seroprevalence estimates. Low sensitivity and specificity can result in overestimation or underestimation of seroprevalence, respectively, thereby potentially compromising the validity of public health decisions based on this data. We mitigated these test imperfections by appropriately adjusting the seroprevalence estimates.

Population immunity for measles significantly increased over the 12-year period in Kilifi HDSS suggesting ongoing improvement in KEPI immunisation program performance over time. The program continues to heavily rely on SIAs for boosting immunity, as evidenced by increased seroprevalence in older age groups following repeated SIAs, particularly in surveys conducted after these campaigns. Efforts should be made to improve both the timing and the coverage of the 1^st^ and 2^nd^ doses to reduce the reliance on frequent SIAs in later years of life. The implementation of rubella vaccination has demonstrated a positive impact on population immunity in children. It is crucial to sustain this immunity to prevent potential gaps in older age groups, as such gaps could lead to an elevated risk of CRS in infants.

## Data Availability

All data produced in the present study are available upon reasonable request to the authors

## DECLARATIONS

## Acknowledgements

This research is funded by an MRC/DFID African Research Leader Fellowship (MR/S005293/1: IMOA, CNM, RC, RO, and AS) and the Bill & Melinda Gates Foundation (INV-039626: AS and EWK). IMOA and JAGS have received grants from the Gavi, the Vaccine Alliance. JAGS is funded by a Wellcome Trust Senior Research Fellowship (214320) and the NIHR Health Protection Research Unit in Immunisation. We thank Dr. Laura Hammitt, Ms. Angela Karani, the residents of the Kilifi Health and Demographic Surveillance System and the dedicated team of fieldworkers, administrative staff, clinicians, and laboratory staff who worked on this study. We acknowledge the continued collaboration of colleagues and the leadership in the Kilifi County Department of Health. This report is published with the permission of the Director of the Kenya Medical Research Institute This research was partly funded by the National Institute for Health Research (NIHR) using UK aid from the UK Government to support global health research. The views expressed in this publication are those of the author(s) and not necessarily those of the NIHR or the UK Department of Health and Social Care (NIHR Global Health Research Unit on Mucosal Pathogens: JO). Wellcome Trust (208812/Z/17/Z: SFlasche).

## Authors’ contribution

IMOA and SF conceived this study. JO, EWK and JAGS contributed to the study design. DA, AS and BK contributed to participant recruitment and data collection. RC and RO generated the serology data and contributed to the analysis assisted by TJ, GS, PGMvG and FRMvdK. CNM conducted the analysis with the guidance of IMOA, SF and JO. All authors contributed in interpretation of findings, writing and critical revision of the manuscript. All authors have read and approved the final manuscript.

