## Supplemental files for "Seroprevalence of Immunoglobulin G against measles and rubella over a 12-year period (2009 – 2021) in Kilifi, Kenya and the impact of the Measles-Rubella (MR) Vaccine campaign of 2016"

Table s1: Mid-year population estimates in KHDSS. The average population across the years was used as the standard population.

| Age_yrs | 2009 | 2010 | 2011 | 2012 | 2013 | 2014 | 2015 | 2016 | 2017 | 2018 | 2019 | 2020 | 2021 |
| --- | --- | --- | --- | --- | --- | --- | --- | --- | --- | --- | --- | --- | --- |
| 0 | 9620 | 9590 | 10738 | 10191 | 10418 | 10779 | 10372 | 9487 | 9915 | 9285 | 9348 | 7866 | 8225 |
| 1 | 9688 | 9930 | 8835 | 9950 | 9284 | 9186 | 9743 | 9158 | 9092 | 9196 | 9295 | 8878 | 8275 |
| 2 | 9078 | 9769 | 9818 | 8853 | 9975 | 9180 | 8998 | 9398 | 9218 | 9163 | 9187 | 9293 | 9003 |
| 3 | 9447 | 9098 | 9624 | 9787 | 8903 | 9810 | 9057 | 9001 | 9440 | 9128 | 9057 | 8972 | 8985 |
| 4 | 8515 | 9327 | 8997 | 9617 | 9805 | 8849 | 9747 | 9068 | 8930 | 9436 | 9087 | 8812 | 8961 |
| 5 | 8445 | 8448 | 9295 | 9012 | 9709 | 9759 | 8715 | 9650 | 9038 | 8992 | 9478 | 9222 | 8817 |
| 6 | 8703 | 8442 | 8341 | 9272 | 8962 | 9589 | 9652 | 8759 | 9798 | 9119 | 9031 | 9458 | 9005 |
| 7 | 8062 | 8641 | 8442 | 8311 | 9301 | 8998 | 9490 | 9726 | 8871 | 9766 | 9140 | 8939 | 9305 |
| 8 | 8257 | 8000 | 8634 | 8374 | 8348 | 9199 | 8995 | 9635 | 9622 | 8794 | 9759 | 9098 | 8877 |
| 9 | 8153 | 8173 | 7899 | 8514 | 8324 | 8317 | 9153 | 8988 | 9545 | 9735 | 8820 | 9452 | 8985 |
| 10 | 7633 | 8054 | 8120 | 7898 | 8505 | 8297 | 8228 | 9156 | 9014 | 9604 | 9747 | 9151 | 9592 |
| 11 | 7093 | 7665 | 8138 | 8216 | 7900 | 8452 | 8202 | 8210 | 9157 | 9004 | 9564 | 9428 | 8668 |
| 12 | 7212 | 7028 | 7653 | 8139 | 8110 | 7883 | 8368 | 8274 | 8119 | 9177 | 8916 | 9459 | 9560 |
| 13 | 6983 | 7139 | 7043 | 7602 | 8118 | 7946 | 7762 | 8326 | 8131 | 8103 | 9064 | 8942 | 9395 |
| 14 | 6448 | 6863 | 7026 | 6851 | 7331 | 7919 | 7766 | 7557 | 8133 | 8005 | 7833 | 8703 | 8592 |

| Age(yrs) | Average population KHDSS  (2009-2021) | Malaria survey  (2009,2011,2013) | PCVIS survey (2015,2017,2019) | COVID19 survey  (2021) | Study population |
| --- | --- | --- | --- | --- | --- |
| 0 | 9680 | 34 | 99 | 8 | 141 |
| 1 | 9270 | 93 | 122 | 23 | 238 |
| 2 | 9303 | 74 | 117 | 23 | 214 |
| 3 | 9255 | 81 | 139 | 23 | 243 |
| 4 | 9165 | 84 | 128 | 15 | 227 |
| 5 | 9122 | 83 | 135 | 17 | 235 |
| 6 | 9087 | 116 | 124 | 19 | 259 |
| 7 | 8999 | 95 | 143 | 24 | 262 |
| 8 | 8892 | 97 | 78 | 18 | 193 |
| 9 | 8774 | 86 | 75 | 25 | 186 |
| 10 | 8692 | 107 | 24 | 27 | 158 |
| 11 | 8438 | 51 | 30 | 15 | 96 |
| 12 | 8300 | 47 | 44 | 19 | 110 |
| 13 | 8043 | 20 | 44 | 17 | 81 |
| 14 | 7617 | 11 | 15 | 17 | 43 |
| Sex |  |  |  |  |  |
| Male | 67016 | 552 | **671** | **158** | 1381 |
| Female | 65620 | 527 | **646** | **132** | 1305 |

Table s2: This table presents mid-year population estimates averaged over the study period in KHDSS, along with total population figures for the Malaria, PCVIS, and COVID-19 surveys. All samples from the PCVIS and COVID-19 surveys were utilized in our study. Population estimates for the malaria survey reflect the figures used in the study. The numbers for the original Malaria study were not available.

Table s3: Standardized GMCs for measles and by age group and year using a reference population to facilitate comparison across years.

| Survey year | 2009 | | | 2011 | | | 2013 | | | 2015 | | | 2017 | | | 2019 | | | 2021 | | |
| --- | --- | --- | --- | --- | --- | --- | --- | --- | --- | --- | --- | --- | --- | --- | --- | --- | --- | --- | --- | --- | --- |
| Measles | N | GMC [95% CI] | | N | GMC [95% CI] | | N | GMC [95% CI] | | N | GMC [95% CI] | | N | GMC [95% CI] | | N | GMC [95% CI] | | N | GMC [95% CI] | |
| Age in years |  |  |  |  |  |  |  |  |  |  |  |  |  |  |  |  |  |  |  |  |  |
| <9m | 4 | 0 | [0-0] | 8 | 0 | [0-0] | 3 | 0 | [0-0.04] | 14 | 0 | [0-0.01] | 15 | 0 | [0-0] | 20 | 0 | [0-0] | 5 | 0 | [0-0.11] |
| 9-17m | 26 | 0.06 | [0.02-0.19] | 9 | 0.01 | [0-0.23] | 19 | 0.02 | [0.01-0.05] | 35 | 0.01 | [0-0.02] | 36 | 0.01 | [0-0.02] | 46 | 0.02 | [0.01-0.04] | 15 | 0.03 | [0.01-0.12] |
| 18-29m | 34 | 0.04 | [0.01-0.1] | 24 | 0.12 | [0.07-0.19] | 38 | 0.08 | [0.06-0.12] | 31 | 0.08 | [0.03-0.19] | 34 | 0.1 | [0.06-0.17] | 56 | 0.06 | [0.03-0.1] | 20 | 0.14 | [0.1-0.2] |
| 30-59m | 80 | 0.16 | [0.12-0.21] | 54 | 0.31 | [0.24-0.39] | 67 | 0.35 | [0.29-0.44] | 97 | 0.15 | [0.1-0.21] | 100 | 0.27 | [0.21-0.36] | 121 | 0.14 | [0.11-0.18] | 52 | 0.14 | [0.08-0.26] |
| 5-9yrs | 197 | 0.18 | [0.15-0.21] | 137 | 0.17 | [0.14-0.2] | 143 | 0.26 | [0.21-0.33] | 152 | 0.22 | [0.17-0.28] | 182 | 0.22 | [0.19-0.26] | 221 | 0.21 | [0.18-0.26] | 103 | 0.20 | [0.14-0.27] |
| 10-14yrs | 24 | 0.08 | [0.04-0.15] | 76 | 0.08 | [0.06-0.11] | 136 | 0.09 | [0.07-0.12] | 47 | 0.11 | [0.07-0.17] | 54 | 0.2 | [0.16-0.27] | 56 | 0.18 | [0.14-0.23] | 95 | 0.18 | [0.15-0.22] |
| Total | 365 | 0.52 | [0.34-0.86] | 308 | 0.69 | [0.51-1.12] | 406 | 0.8 | [0.64-1.1] | 376 | 0.57 | [0.37-0.88] | 421 | 0.8 | [0.62-1.08] | 520 | 0.61 | [0.47-0.81] | 290 | 0.69 | [0.48-1.18] |
| P_value |  | <0.001 | |  | <0.001 | |  | <0.001 | |  | <0.001 | |  | <0.001 | |  | <0.001 | |  | 0.001 | |

Table s4 :Standardized GMCs for rubella by age group and year using a reference population to facilitate comparison across years.

| Survey  year | 2009 | | | 2011 | | | 2013 | | | 2015 | | | 2017 | | | 2019 | | | | 2021 | | |
| --- | --- | --- | --- | --- | --- | --- | --- | --- | --- | --- | --- | --- | --- | --- | --- | --- | --- | --- | --- | --- | --- | --- |
| Rubella | N | GMC [95% CI] | | N | GMC [95% CI] | | N | GMC [95% CI] | | N | GMC [95% CI] | | N | GMC [95% CI] | | N | GMC [95% CI] | | N | | GMC [95% CI] | |
| Age in  years |  |  |  |  |  |  |  |  |  |  |  |  |  |  |  |  |  |  |  | |  |  |
| <9m | 4 | 0.06 | [0.01-0.4] | 8 | 0.02 | [0.01-0.03] | 3 | 0.13 | [0.01-2.18] | 14 | 0.05 | [0.01-0.32] | 15 | 0.08 | [0.03-0.24] | 20 | 0.06 | [0.02-0.16] | 5 | | 1.83 | [0.23-14.38] |
| 9-17m | 26 | 0.01 | [0-0.01] | 9 | 0.04 | [0-0.89] | 19 | 0.01 | [0-0.02] | 35 | 0.02 | [0-0.05] | 36 | 0.37 | [0.1-1.43] | 46 | 1.75 | [0.77-3.98] | 15 | | 3.08 | [0.59-16.1] |
| 18-29m | 34 | 0.01 | [0-0.02] | 24 | 0.02 | [0.01-0.08] | 38 | 0.01 | [0-0.01] | 31 | 0.02 | [0-0.06] | 34 | 0.96 | [0.24-3.84] | 56 | 4.87 | [3.07-7.72] | 20 | | 14.02 | [10.91-18.06] |
| 30-59m | 80 | 0.1 | [0.05-0.22] | 54 | 0.02 | [0.01-0.05] | 67 | 0.03 | [0.02-0.06] | 97 | 0.14 | [0.06-0.32] | 100 | 6.16 | [3.05-12.43] | 121 | 9.63 | [6.24-14.84] | 52 | | 9.35 | [4.59-18.94] |
| 5-9yrs | 197 | 1.52 | [0.89-2.6] | 137 | 2.7 | [1.3-5.65] | 143 | 0.32 | [0.16-0.64] | 152 | 0.95 | [0.45-2.03] | 182 | 32.9 | [22.4-48.4] | 221 | 16.35 | [11.13-23.99] | 103 | | 21.29 | [12.72-35.54] |
| 10-14yrs | 24 | 6.03 | [1.17-31.05] | 76 | 11.32 | [5.27-24.41] | 136 | 7.85 | [4.32-14.34] | 47 | 3.53 | [0.96-13] | 54 | 52.7 | [32.26-85.83] | 56 | 39.57 | [24.02-65.44] | 95 | | 46.27 | [33.48-63.61] |
| Total | 365 | 7.73 | [2.12-34.3] | 308 | 14.12 | [6.6-31.11] | 406 | 8.35 | [4.51-17.25] | 376 | 4.71 | [1.48-15.78] | 421 | 93.13 | [58.08-152.17] | 520 | 72.23 | [45.25-116.13] | 290 | | 95.84 | [62.52-166.63] |
| P_value |  | <0.001 | |  | <0.001 | |  | <0.001 | |  | <0.001 | |  | <0.001 | |  | <0.001 | |  | | 0.01 | |

Table s5; Standardized age-specific measles seroprevalence estimates and Geometric Mean concentrations (GMCs) in adults. Seroprevalence estimates were also adjusted for test performance using Bayesian modelling

| Survey year | 2021 | | | | | | |
| --- | --- | --- | --- | --- | --- | --- | --- |
| Measles | n | % [95% CI] | | P_value | GMC [95% CI] | | P_value |
| Age in years |  |  |  |  |  |  |  |
| 15_19 | 56 | 97 | [87-100] | 0.41 | 0.05 | [0.04-0.07] | <0.001 |
| 20_24 | 51 | 96 | [85-100] |  | 0.05 | [0.03-0.08] |  |
| 25_29 | 47 | 97 | [88-100] |  | 0.05 | [0.03-0.07] |  |
| 30_34 | 46 | 99 | [94-100] |  | 0.06 | [0.04-0.09] |  |
| 35_39 | 54 | 99 | [91-100] |  | 0.04 | [0.03-0.06] |  |
| 40_44 | 44 | 97 | [86-100] |  | 0.05 | [0.03-0.08] |  |
| 45-49 | 56 | 99 | [95-100] |  | 0.04 | [0.03-0.06] |  |
| 50_54 | 48 | 99 | [95-100] |  | 0.05 | [0.04-0.07] |  |
| 55_59 | 50 | 98 | [88-100] |  | 0.04 | [0.03-0.07] |  |
| 60_64 | 48 | 99 | [96-100] |  | 0.03 | [0.02-0.03] |  |
| 65+ | 55 | 99 | [95-100] |  | 0.1 | [0.08-0.14] |  |
| Total | 555 | 99 | [95-99] |  | 0.56 | [0.40-0.82] |  |

Table s6: Standardized age-specific rubella seroprevalence estimates and Geometric Mean concentrations (GMCs) in adults. Seroprevalence estimates were also adjusted for test performance using Bayesian modelling.

| Survey year | 2021 | | | | | | |
| --- | --- | --- | --- | --- | --- | --- | --- |
| Rubella | n | % [95% CI] |  | P_value | GMC [95% CI] |  | P_value |
| 15_19 | 56 | 99 | [94-100] | 0.91 | 18.83 | [15.28-23.02] | 0.14 |
| 20_24 | 51 | 90 | [80-98] |  | 5.27 | [2.53-10.99] |  |
| 25_29 | 47 | 88 | [75-97] |  | 3.11 | [1.34-7.2] |  |
| 30_34 | 46 | 86 | [74-96] |  | 2.18 | [0.82-5.73] |  |
| 35_39 | 54 | 95 | [84-99] |  | 2.41 | [1.23-4.68] |  |
| 40_44 | 44 | 93 | [81-99] |  | 2.88 | [1.49-5.54] |  |
| 45-49 | 56 | 93 | [84-99] |  | 1.86 | [0.98-3.52] |  |
| 50_54 | 48 | 95 | [87-100] |  | 2.49 | [1.77-3.49] |  |
| 55_59 | 50 | 92 | [82-99] |  | 1.36 | [0.72-2.6] |  |
| 60_64 | 48 | 92 | [80-99] |  | 1.37 | [0.76-2.46] |  |
| 65+ | 55 | 97 | [90-100] |  | 5.08 | [3.22-8.02] |  |
| Total | 555 | 92 | [89-96] |  | 46.84 | [30.14-77.25] |  |

Table s7: Measles and rubella seroprevalence and IgG GMCs before and after the 2016 MR vaccine campaign. Seroprevalence estimates were calculated by using multilevel regression and poststratification (MLRP) year*s* and subsequently adjusted for assay sensitivity and specificity

| Survey year | 2015 | | | | | 2017 | | | | | P-value % | P-value GMCs |
| --- | --- | --- | --- | --- | --- | --- | --- | --- | --- | --- | --- | --- |
|  |  | (10-15 months before the campaign) | | | | (7-12 months after the campaign) | | | | |  |  |
| Measles | N | % [95% CI] | | GMC [95% CI] | | N | % [95% CI] | | GMC [95% CI] | |  |  |
| Age in years |  |  |  |  |  |  |  |  |  |  |  |  |
| 9-17m | 35 | 71 | [51-86] | 0.01 | [0-0.02] | 2 | 96 | [68-100] | 0.09 | [0-0.11] | 0.81 | 0.03 |
| 18-29m | 31 | 95 | [82-100] | 0.08 | [0.03-0.19] | 32 | 98 | [89-100] | 0.09 | [0.05-0.16] | 0.58 | 0.7 |
| 30-59m | 97 | 96 | [88-100] | 0.14 | [0.1-0.21] | 100 | 98 | [94-100] | 0.27 | [0.2-0.36] | 0.19 | 0.01 |
| 5-9yrs | 152 | 97 | [92-100] | 0.22 | [0.17-0.28] | 182 | 98 | [94-100] | 0.23 | [0.19-0.27] | 0.53 | 0.86 |
| 10-14yrs | 36 | 94 | [80-100] | 0.13 | [0.07-0.21] | 54 | 97 | [89-100] | 0.21 | [0.16-0.27] | 0.57 | 0.09 |
| Total | 351 | 92 | [85-94] | 0.58 | [0.37-0.91] | 370 | 96 | [89-98] | 0.89 | [0.60-1.17] | 0.01 | 0.001 |
| Rubella |  |  |  |  |  |  |  |  |  |  |  |  |
| 9-17m | 35 | 16 | [03-34] | 0.02 | [0-0.05] | 2 | 55 | [09-94] | 0.08 | [0-0.28] | 0.34 | 0.001 |
| 18-29m | 31 | 15 | [03-33] | 0.02 | [0-0.06] | 32 | 66 | [46-82] | 0.93 | [0.22-3.94] | <0.001 | <0.001 |
| 30-59m | 97 | 29 | [14-42] | 0.14 | [0.06-0.32] | 100 | 81 | [71-92] | 6.01 | [2.98-12.14] | <0.001 | <0.001 |
| 5-9yrs | 152 | 51 | [39-61] | 0.97 | [0.45-2.06] | 182 | 93 | [85-98] | 33.4 | [22.8-49.14] | <0.000 | <0.001 |
| 10-14yrs | 36 | 65 | [46-81] | 3.03 | [0.66-13.83] | 54 | 96 | [86-99] | 53.05 | [32.5-86.48] | <0.001 | <0.001 |
| Total | 351 | 45 | [35-53] | 4.18 | [1.17-16.32] | 370 | 82 | [77-88] | 93.47 | [58.5-151.98] | <0.001 | <0.001 |

Table s8: Measles and Rubella Seroprevalence and IgG GMCs in children <9m of age in the different years

| Survey year | 2009-2021 | | | | |
| --- | --- | --- | --- | --- | --- |
| Measles | N | % | [95%CI] | GMC | [95% CI] |
| Age(months) |  |  |  |  |  |
| 1 | 1 | 100 | [21-100] | 2.61 | Na |
| 3 | 1 | 0 | [0-79] | 0.001 | Na |
| 4 | 11 | 27 | [09-57] | 0.06 | [0.02-0.01] |
| 5 | 8 | 0 | [0-32] | 0.01 | [0.01-0.04] |
| 6 | 14 | 21 | [08-47] | 0.02 | [0.01-0.05] |
| 7 | 11 | 9 | [02-38] | 0.02 | [0.01-0.06] |
| 8 | 16 | 6 | [01-28] | 0.01 | [0.01-0.01] |
| 9 | 7 | 14 | [02-51] | 0.01 | [0.01-0.05] |
| Rubella |  |  |  |  |  |
| 1 | 1 | 100 | [21-100] | 177 | Na |
| 3 | 1 | 0 | [0-79] | 0.01 | Na |
| 4 | 11 | 36 | [15-64] | 4.31 | [0.76-24.5] |
| 5 | 8 | 0 | [0-32] | 2.55 | [0.75-8.64] |
| 6 | 14 | 14 | [4-40] | 1.93 | [0.49-7.53] |
| 7 | 11 | 9 | [2-38] | 1.42 | [0.61-3.34] |
| 8 | 16 | 0 | [0-19] | 0.38 | [0.17-0.85] |
| 9 | 7 | 14 | [3-51] | 0.4 | [0.02-7.79] |

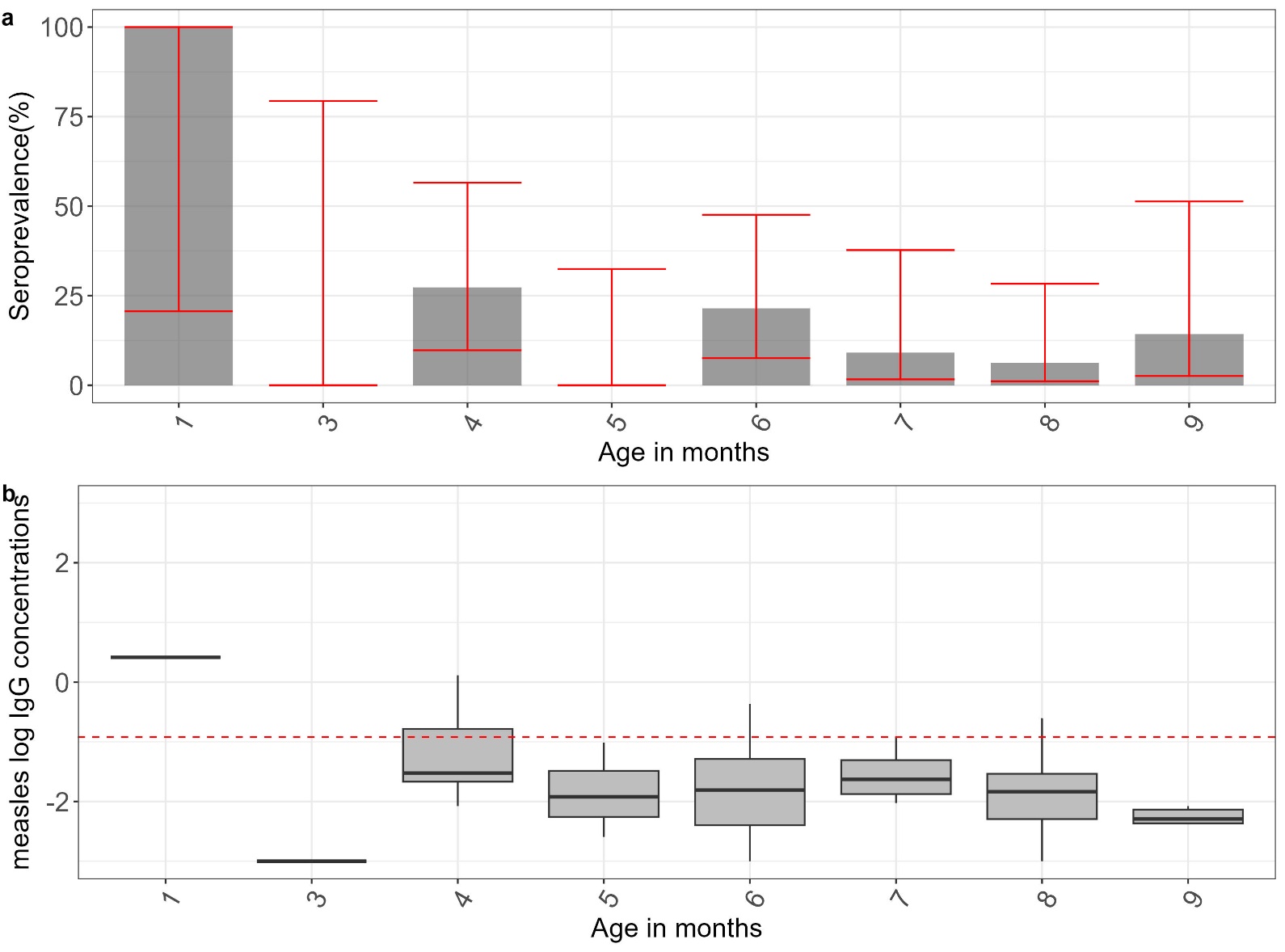

Figure s1.Ia shows measles seroprevalence estimates in infants. The red error bars indicate 95% credible intervals. 1b shows the distribution of antibodies with the median indicated by the line in the box and a whisker plot. The red line in the lower figure is the log of the protective threshold for measles of ≥0.12 IU/ml

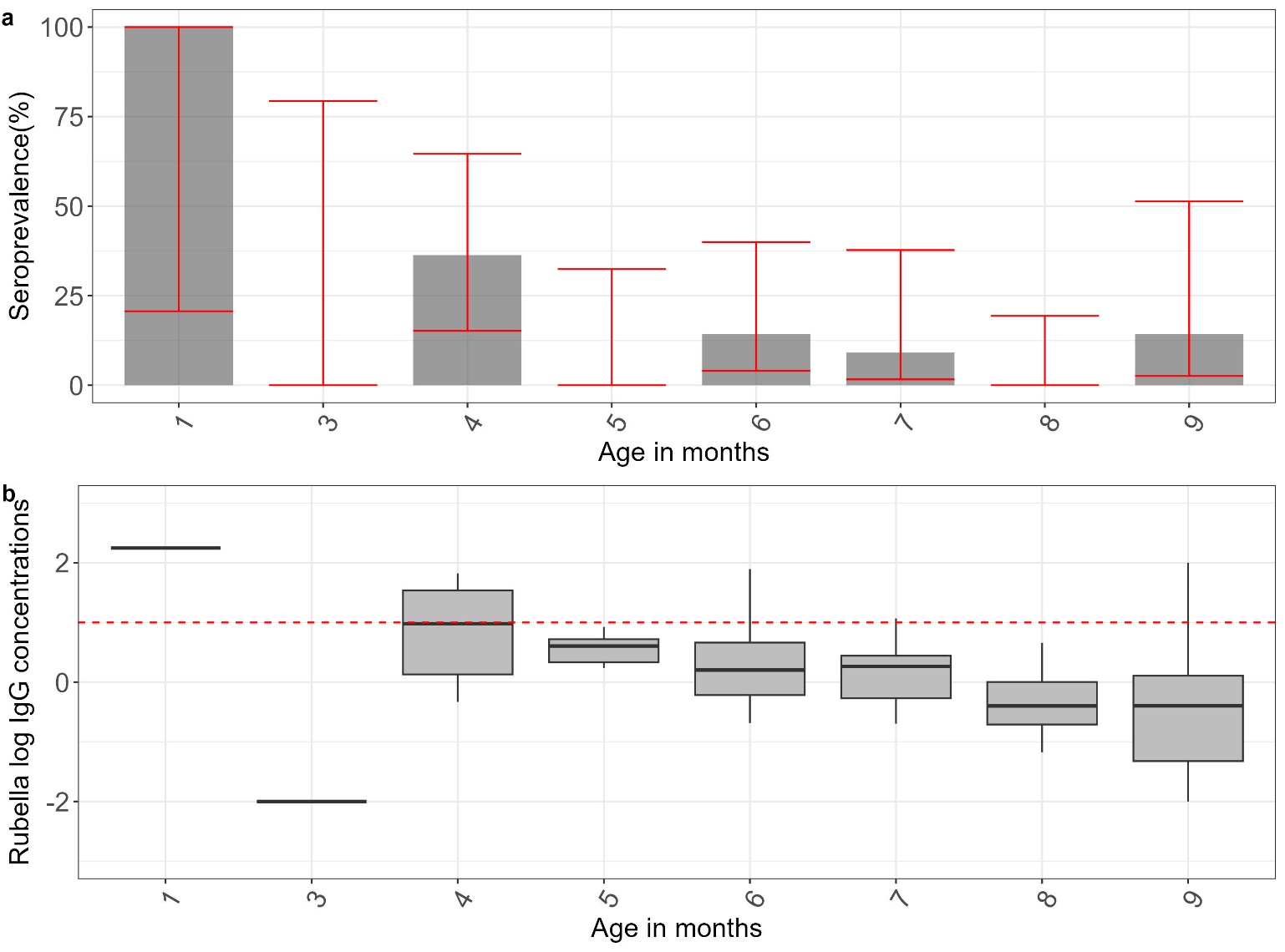

Figure s2.2a shows rubella seroprevalence estimates in infants. The red error bars indicate 95% credible intervals. 1b shows the distribution of antibodies with the median indicated by the line in the box and a whisker plot. The red line in the lower figure is the log of the protective threshold for rubella of ≥10 IU/ml
